# Prospective Validation of a Deep Learning Model to Detect Structural Heart Disease from Apple Watch ECGs: The WATCH-SHD Study

**DOI:** 10.64898/2026.08.13.26360395

**Authors:** Arya Aminorroaya, Sumukh Vasisht Shankar, Madeleine Carter, Mariam Khan, Lovedeep S Dhingra, Akshay Khunte, Philip M Croon, Bernardo Lombo, Robert L McNamara, Evangelos K Oikonomou, Aline F Pedroso, Rohan Khera

## Abstract

**Importance:** Consumer wearables such as the Apple Watch can record single-lead electrocardiograms (ECGs) but are used mainly to detect rhythm disorders. Artificial intelligence–enhanced ECG (AI-ECG) could extend these real-world recordings for detecting structural heart disease (SHD), yet prospective validation remains limited.

**Objective:** To prospectively validate a previously developed, noise-adapted AI-ECG model for detecting severe SHD from single-lead Apple Watch ECGs.

**Design:** Prospective cohort study.

**Setting:** Yale New Haven Hospital echocardiography laboratory.

**Participants:** Adults aged ≥18 years undergoing outpatient transthoracic echocardiography (TTE) as part of routine clinical care.

**Exposure:** A 30-second, single-lead Apple Watch ECG recorded during the TTE visit and processed through an end-to-end, HIPAA-compliant platform for real-time AI-ECG inference.

**Main Outcomes and Measures:** The primary outcome was discrimination for TTE-defined severe SHD—a composite of left ventricular systolic dysfunction (left ventricular ejection fraction <40%), severe left-sided valvular disease, and/or severe left ventricular hypertrophy—assessed by the area under the receiver operating characteristic curve (AUROC). Secondary measures were sensitivity, specificity, negative predictive value (NPV), and positive predictive value (PPV) at prespecified thresholds, and screening efficiency, assessed by the number needed to test (NNT) under usual-care versus AI-ECG–guided strategies.

**Results:** Among 596 participants with analyzable Apple Watch ECGs (median age, 62 years [IQR, 46–72]; 51.2% women), severe SHD was present in 30 (5.1%). The model discriminated severe SHD well (AUROC, 0.841; 95% CI, 0.761–0.921), with a sensitivity of 76.7% (59.1–88.2), specificity of 83.2% (79.9–86.1), NPV of 98.5% (97.0–99.3), and PPV of 19.7% (13.5–27.8) at the prespecified threshold. An AI-ECG–guided strategy reduced the NNT to identify one case by more than 60% versus usual care across the composite and individual SHD phenotypes.

**Conclusions and Relevance:** In this prospective cohort, a noise-adapted AI-ECG algorithm identified SHD phenotypes from real-world single-lead Apple Watch ECGs and improved screening efficiency. These findings support a potential role for wearable ECG–based screening in the scalable identification of clinically actionable SHD.

**KEY POINTS:** *Question:* Can a noise-adapted AI-ECG model accurately detect severe structural heart disease (SHD) from single-lead Apple Watch ECGs?

*Findings:* In this prospective cohort of 596 adults undergoing outpatient transthoracic echocardiography, AI-ECG applied to Apple Watch ECG recordings discriminated severe SHD well (AUROC, 0.841), with 76.7% sensitivity and 83.2% specificity. An AI-ECG–guided strategy lowered the number needed to test to identify one case by more than 60% versus usual care, across the composite and individual SHD phenotypes.

*Meaning:* These findings provide prospective evidence that wearable single-lead ECG–based screening could enable accessible, scalable identification of clinically actionable SHD.

## INTRODUCTION

Without scalable screening strategies, structural heart disease (SHD) often goes undetected until symptoms or complications develop.^1–8^ Because effective therapies now exist for SHD phenotypes such as left ventricular systolic dysfunction (LVSD) and valvular disease, a delayed diagnosis forfeits the window when early treatment is most beneficial.^1–6^ Yet systematic screening of asymptomatic individuals is rarely implemented, because the confirmatory modalities, chiefly echocardiography, are costly, resource-intensive, and impractical to deploy at the population scale.^9–11^ A scalable, broadly accessible screening tool is therefore needed.

The widespread adoption of wearable devices such as Apple Watch offers an opportunity to extend cardiovascular screening beyond clinical settings.^12,13^ Although these devices are used mainly for rhythm monitoring,^14,15^ artificial intelligence (AI) can detect the subtle electrical signatures that structural abnormalities leave on electrocardiograms (ECGs), including single-lead recordings.^16–20^ Existing AI-ECG models, however, have been trained almost exclusively on high-fidelity clinical ECGs, raising questions about how well they generalize to the noisier signals of consumer-grade devices.^19,20^ Furthermore, efforts to apply single-lead ECGs for SHD detection have typically focused on isolated phenotypes, most often LVSD.^20–23^ Although technically feasible, this narrow focus limits clinical utility: any single SHD phenotype is uncommon in the community, yielding a low positive predictive value (PPV), a critical metric for screening.^17–19^

We addressed these gaps by prospectively evaluating a previously developed, noise-adapted deep learning model that detects a composite of clinically actionable severe SHD—LVSD, severe left-sided valvular disease, and severe left ventricular hypertrophy (LVH)—from lead I of clinical ECGs. The model was trained and externally validated on clinical single-lead ECGs derived from health-system data.^19^ Here, we tested its performance on real-world single-lead Apple Watch ECGs in patients undergoing transthoracic echocardiography (TTE).

## METHODS

### Study Design

WATCH-SHD was a prospective validation study at the Yale New Haven Hospital (YNHH) echocardiography laboratory. During the same visit as a clinically indicated outpatient TTE, participants recorded a single-lead Apple Watch ECG (**Figure 1**). Each recording was stored and then linked to the structured TTE report from the electronic health record (EHR) through a unique participant identifier.

**Figure 1.**
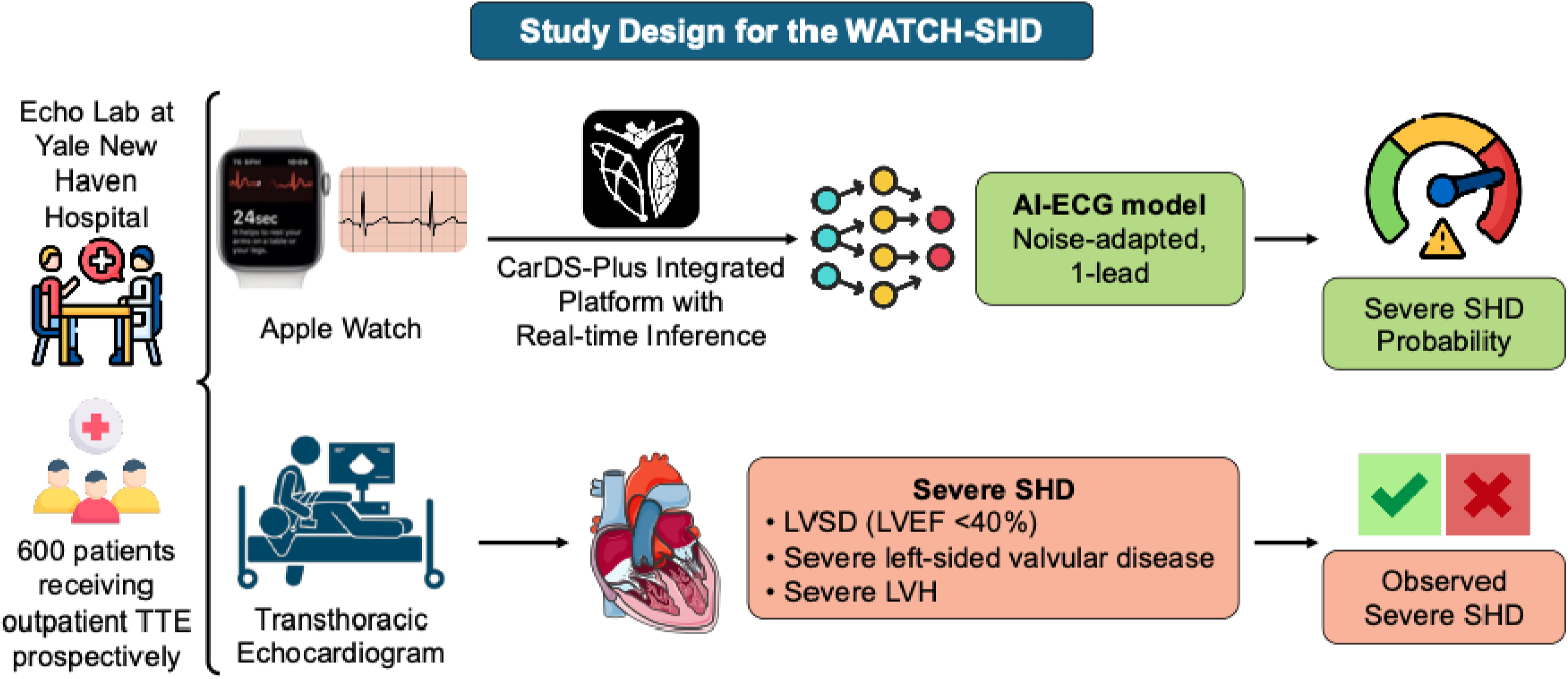
Study Design for the WATCH-SHD. **Abbreviations:** AI-ECG, artificial intelligence-enhanced electrocardiogram; LVEF, left ventricular ejection fraction; LVH, left ventricular hypertrophy; LVSD, left ventricular systolic dysfunction; SHD, structural heart disease; TTE, transthoracic echocardiogram.

### Study Population

Between June 2024 and January 2025, we enrolled adults aged ≥18 years scheduled for outpatient TTE at the YNHH echocardiography laboratory. We excluded patients with a permanent pacemaker or implantable cardioverter-defibrillator, pregnant individuals, and those with cognitive impairment or an inability to communicate in English. All participants provided written informed consent.

### Study Covariates

Severe SHD was a composite echocardiographic diagnosis of LVSD, severe left-sided valvular disease, and/or severe LVH. We also defined a broader SHD category that additionally counted moderate (not only severe) left-sided valvular disease alongside LVSD and/or severe LVH. LVSD was a left ventricular ejection fraction (LVEF) below 40%; left-sided valvular disease comprised aortic or mitral stenosis or regurgitation; and severe LVH was an interventricular septal thickness in diastole (IVSd) above 15 mm accompanied by moderate or severe left ventricular diastolic dysfunction. All measurements followed American Society of Echocardiography guidelines.^11^ LVEF was quantified by three-dimensional echocardiography or the Simpson biplane method, when available, or by visual estimation. IVSd was measured quantitatively, whereas valvular and diastolic dysfunction severity were graded by the interpreting cardiologist using guideline-recommended criteria.^11^

We used the EHR to ascertain key clinical covariates: hypertension, type 2 diabetes (T2D), atherosclerotic cardiovascular disease (ASCVD), and heart failure (HF). ASCVD was defined by a history of coronary artery disease (CAD), ischemic stroke, transient ischemic attack (TIA), or peripheral artery disease (PAD). All covariates were identified from International Classification of Diseases, Tenth Revision, Clinical Modification (ICD-10-CM) codes, as previously described.^24–26^

### Apple Watch ECG Acquisition

Participants recorded a 30-second, single-lead ECG on an Apple Watch Series 4 worn on the left wrist, with the right index finger placed on the digital crown. A 41-mm or 45-mm case was selected by wrist size, per manufacturer guidance.

### AI-ECG Model and Prospective Inference

We applied a previously developed, noise-adapted, ensemble deep learning model that detects severe SHD from lead I of clinical ECGs.^19^ In WATCH-SHD, single-lead ECGs were captured on the Apple Watch through CarDS-Plus, an end-to-end, Health Insurance Portability and Accountability Act (HIPAA)–compliant platform we developed previously.^27^ The CarDS-Plus platform enabled ECG recording, secure transmission, and real-time inference during the study visit in the YNHH echocardiography laboratory. Because Apple Watch tracings span 30 seconds—versus the 10 seconds of a standard clinical ECG—each recording was divided into five overlapping 10-second segments (0–10, 5–15, 10–20, 15–25, and 20–30 seconds).

Waveforms were downsampled from 512 to 500 Hz and rescaled by a factor of 500 to match the development ECGs. The ensemble model was applied to each segment, and the median of the five predicted probabilities served as the recording-level output.

### Study Outcomes

The primary outcome was discrimination for TTE-defined composite severe SHD, measured by the area under the receiver operating characteristic curve (AUROC). We also reported threshold-based metrics—sensitivity, specificity, negative predictive value (NPV), and PPV—for severe SHD, for the broader SHD composite, and for individual phenotypes. To gauge screening efficiency, we computed the number needed to test (NNT) to identify one case under usual care and under an AI-ECG–guided strategy. Threshold-based metrics were computed using prespecified, label-specific cut-points set to achieve 90% specificity in the internal validation set of the original development study; each outcome was scored with its corresponding label-specific ensemble model.^19^

### Sample Size Calculation

The sample size was set a priori for the primary endpoint of severe SHD. Assuming an AUROC of 0.91,^19^ an event prevalence of 4.4% from preliminary data, and targeting 80% power at a two-sided α of 0.05, we sized the study so that the lower bound of the 95% confidence interval (CI) for the AUROC would exceed 0.75. Allowing a conservative 10% loss to technical or acquisition failures, we estimated that 600 participants would be required.^28,29^

### Statistical Analysis

Continuous variables were reported as medians with interquartile ranges (IQRs) and categorical variables as counts with percentages. Model performance was summarized by AUROC, sensitivity, specificity, NPV, and PPV. We computed 95% CIs for the AUROC by the DeLong method, and for sensitivity, specificity, NPV, and PPV by the Wilson score method.^30^ The NNT under usual care was the reciprocal of outcome prevalence, whereas the NNT under AI-ECG was the reciprocal of the model’s PPV at the prespecified threshold; we then reported the relative NNT reduction. Performance for detecting severe SHD was also summarized across key demographic and clinical subgroups. Analyses were executed using Python 3.11.2, with two-sided tests at α = 0.05. The Yale Institutional Review Board approved the study protocol (#2000035532).

## RESULTS

### Study Population

Of 600 adults enrolled for outpatient TTE, 596 (99.3%) recorded an analyzable 30-second, single-lead Apple Watch ECG and formed the analysis cohort (**Table 1**); the remaining four could not complete a recording before their visit ended. The median age was 62 years (IQR, 46–72), and 305 (51.2%) were women. Self-reported race and ethnicity were White in 371 (65.4%), Black in 103 (18.2%), Hispanic in 55 (9.7%), Asian in 18 (3.2%), Native American in 2 (0.4%), and other in 18 (3.2%). Hypertension was present in 67.6%, T2D in 28.4%, ASCVD in 48.2%, and HF in 27.7%. On TTE, 30 participants (5.1%) met criteria for severe SHD and 97 (16.5%) for the broader SHD composite; individual phenotypes were LVSD (LVEF <40%) in 15 (2.5%), severe valvular disease in 6 (1.0%), moderate or severe valvular disease in 79 (13.4%), and severe LVH in 9 (1.5%).

**Table 1.**
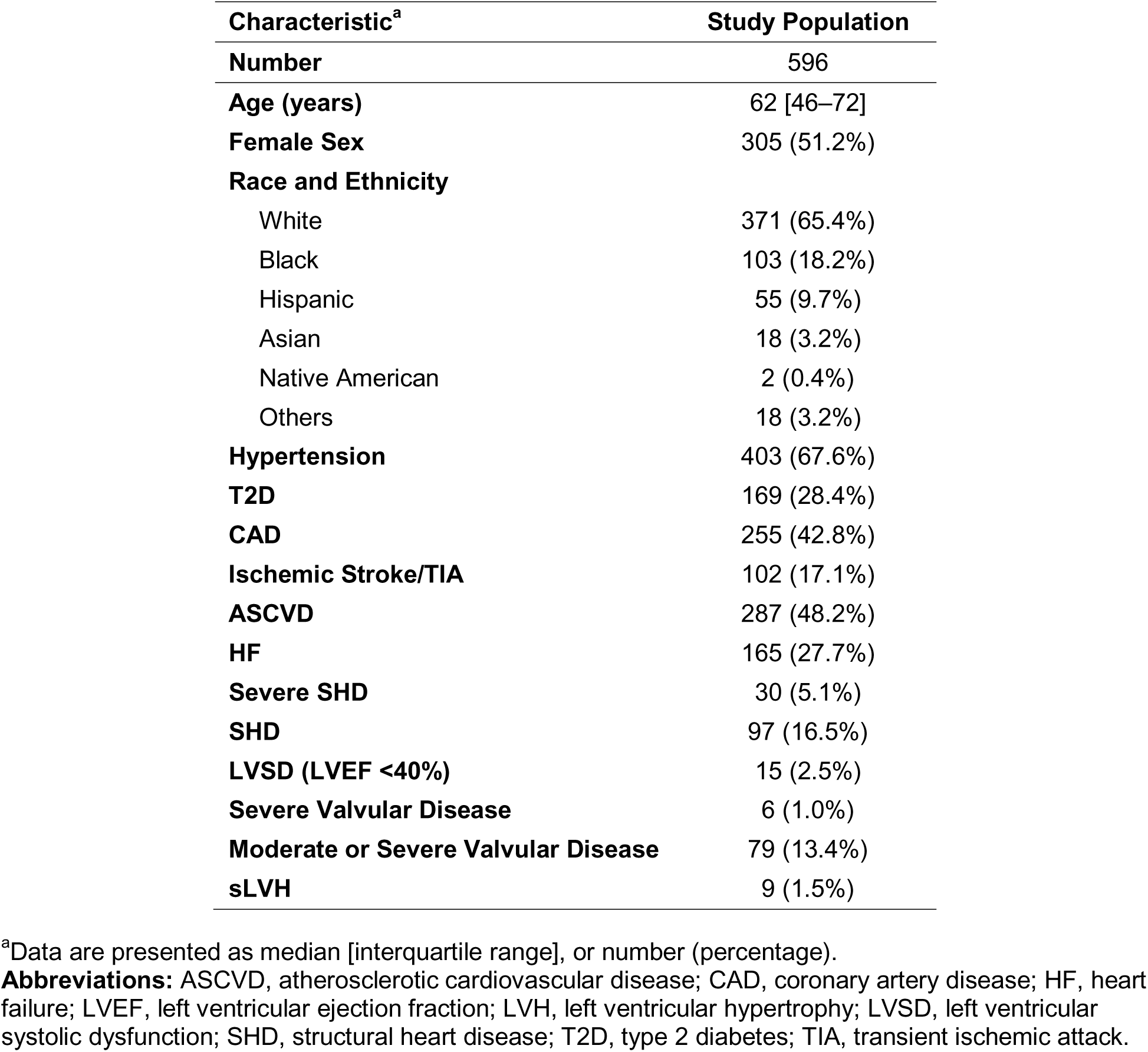
Demographics and Prevalence of Structural Heart Disease in the Study Population.

| **Characteristic<sup>a</sup>** | **Study Population** |
| --- | --- |
| **Number** | 596 |
| **Age (years)** | 62 [46–72] |
| **Female Sex** | 305 (51.2%) |
| **Race and Ethnicity** |  |
| White | 371 (65.4%) |
| Black | 103 (18.2%) |
| Hispanic | 55 (9.7%) |
| Asian | 18 (3.2%) |
| Native American | 2 (0.4%) |
| Others | 18 (3.2%) |
| **Hypertension** | 403 (67.6%) |
| **T2D** | 169 (28.4%) |
| **CAD** | 255 (42.8%) |
| **Ischemic Stroke/TIA** | 102 (17.1%) |
| **ASCVD** | 287 (48.2%) |
| **HF** | 165 (27.7%) |
| **Severe SHD** | 30 (5.1%) |
| **SHD** | 97 (16.5%) |
| **LVSD (LVEF <40%)** | 15 (2.5%) |
| **Severe Valvular Disease** | 6 (1.0%) |
| **Moderate or Severe Valvular Disease** | 79 (13.4%) |
| **sLVH** | 9 (1.5%) |

<sup>a</sup>Data are presented as median [interquartile range], or number (percentage).
**Abbreviations:** ASCVD, atherosclerotic cardiovascular disease; CAD, coronary artery disease; HF, heart failure; LVEF, left ventricular ejection fraction; LVH, left ventricular hypertrophy; LVSD, left ventricular systolic dysfunction; SHD, structural heart disease; T2D, type 2 diabetes; TIA, transient ischemic attack.

### Performance of AI-ECG

The model achieved an AUROC of 0.841 (95% CI, 0.761–0.921) for detecting severe SHD, meeting the primary study outcome, as the lower bound of the 95% CI exceeded 0.75 (**Figure 2A**). At the prespecified threshold, sensitivity was 76.7% (95% CI, 59.1–88.2) and specificity 83.2% (95% CI, 79.9–86.1) (**Table 2)**. At the 5.1% prevalence of severe SHD, NPV was 98.5% (95% CI, 97.0–99.3) and PPV 19.7% (95% CI, 13.5–27.8). Performance was comparable across key demographic and clinical subgroups (**Table 3**).

**Figure 2.**
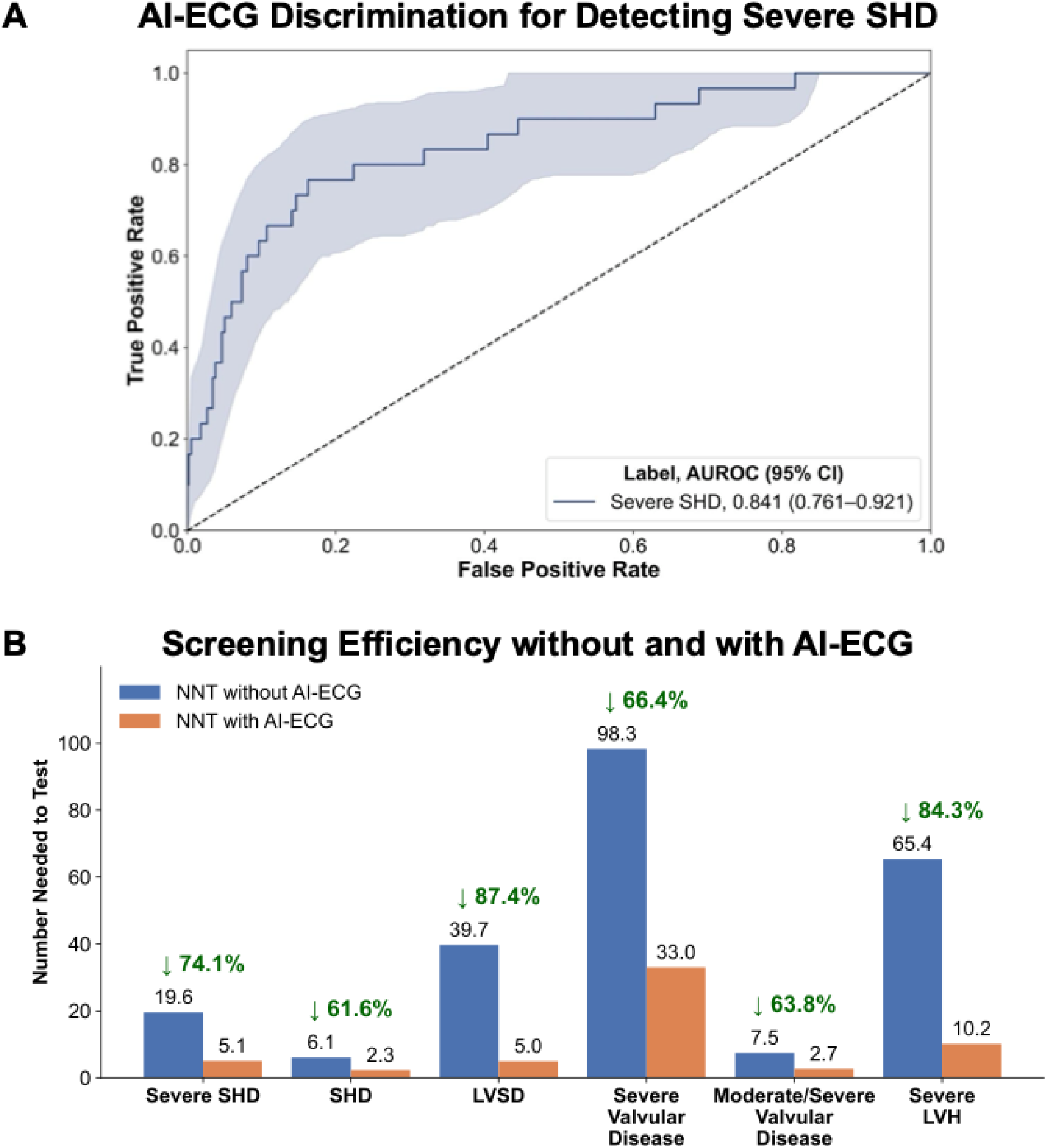
(A) AI-ECG Discrimination for Detecting Severe SHD from Single-lead Apple Watch ECG; (B) Number Needed to Test to Identify One Case from Single-lead Apple Watch ECG without and with AI-ECG Algorithm Across Composite and Individual SHD Phenotypes **Abbreviations:** AI-ECG, artificial intelligence-enhanced electrocardiogram; AUROC, area under the receiver operating characteristic curve; CI, confidence interval; LVH, left ventricular hypertrophy; LVSD, left ventricular systolic dysfunction; NNT, number needed to test; SHD, structural heart disease.

**Table 2.** Performance Measures of the AI-ECG Algorithm Across Composite and Individual Structural Heart Diseases.

| **Label<sup>a</sup>** | **Sensitivity** | **Specificity** | **NPV** | **PPV** | **Prevalence** | **NNT Reduction** |
| --- | --- | --- | --- | --- | --- | --- |
| **Severe SHD<sup>b</sup>** | 76.7% (59.1–88.2) | 83.2% (79.9–86.1) | 98.5% (97.0–99.3) | 19.7% (13.5–27.8) | 30 (5.1%) | 74.1% |
| **SHD<sup>b</sup>** | 52.6% (42.7–62.2) | 86.2% (82.8–88.9) | 90.2% (87.2–92.6) | 42.9% (34.3–51.8) | 97 (16.5%) | 61.6% |
| **LVSD** | 86.7% (62.1–96.3) | 91.0% (88.4–93.1) | 99.6% (98.6–99.9) | 20.0% (12.1–31.3) | 15 (2.5%) | 87.4% |
| **Severe Valvular Disease** | 16.7% (3.0–56.4) | 94.5% (92.4–96.1) | 99.1% (97.9–99.6) | 3.0% (0.5–15.3) | 6 (1.0%) | 66.4% |
| **Moderate or Severe Valvular Disease** | 21.5% (13.9–31.8) | 94.3% (92.0–96.0) | 88.6% (85.7–91.0) | 37.0% (24.5–51.4) | 79 (13.4%) | 63.8% |
| **Severe LVH** | 44.4% (18.9–73.3) | 93.6% (91.3–95.3) | 99.1% (97.9–99.6) | 9.8% (3.9–22.5) | 9 (1.5%) | 84.3% |

<sup>a</sup>Data are presented as point estimate (95% CI), number (percentage), or percentage.
<sup>b</sup>Severe SHD was a composite of LVSD, severe left-sided valvular disease, and/or severe LVH. The broader SHD category additionally counted moderate (not only severe) left-sided valvular disease alongside LVSD and/or severe LVH.
**Abbreviations:** LVH, left ventricular hypertrophy; LVSD, left ventricular systolic dysfunction; NNT, number needed to test; NPV, negative predictive value; PPV, positive predictive value; SHD, structural heart disease.

**Table 3.** Performance Measures of the AI-ECG Algorithm for Detecting Severe SHD Across Key Subgroups.

| **Subgroup<sup>a</sup>** | **Sensitivity** | **Specificity** | **NPV** | **PPV** | **Prevalence** | **NNT Reduction** |
| --- | --- | --- | --- | --- | --- | --- |
| **Overall** | 76.7% (59.1–88.2) | 83.2% (79.9–86.1) | 98.5% (97.0–99.3) | 19.7% (13.5–27.8) | 30 (5.1%) | 74.1% |
| **≥65 years** | 83.3% (55.2–95.3) | 71.6% (65.5–77.0) | 98.8% (95.8–99.7) | 13.0% (7.2–22.3) | 12 (4.8%) | 62.7% |
| **<65 years** | 72.2% (49.1–87.5) | 91.6% (88.1–94.2) | 98.3% (96.2–99.3) | 32.5% (20.1–48.0) | 18 (5.3%) | 83.8% |
| **Female** | 83.3% (55.2–95.3) | 90.3% (86.4–93.2) | 99.2% (97.3–99.8) | 26.3% (15.0–42.0) | 12 (4.0%) | 84.9% |
| **Male** | 72.2% (49.1–87.5) | 75.6% (70.1–80.3) | 97.6% (94.5–99.0) | 16.5% (9.9–26.1) | 18 (6.2%) | 62.0% |
| **White** | 91.7% (64.6–98.5) | 80.8% (76.4–84.6) | 99.7% (98.1–99.9) | 13.9% (8.0–23.2) | 12 (3.3%) | 76.5% |
| **Black** | 73.3% (48.0–89.1) | 83.9% (74.8–90.2) | 94.8% (87.4–98.0) | 44.0% (26.7–62.9) | 15 (14.7%) | 66.6% |
| **Hypertension** | 91.7% (74.2–97.7) | 77.7% (73.2–81.6) | 99.3% (97.6–99.8) | 20.8% (14.1–29.4) | 24 (6.0%) | 71.1% |
| **No Hypertension** | 16.7% (3.0–56.4) | 94.5% (90.2–97.0) | 97.2% (93.6–98.8) | 9.1% (1.6–37.7) | 6 (3.2%) | 65.1% |
| **T2D** | 78.6% (52.4–92.4) | 74.2% (66.8–80.4) | 97.5% (92.8–99.1) | 21.6% (12.5–34.6) | 14 (8.3%) | 61.6% |
| **No T2D** | 75.0% (50.5–89.8) | 86.6% (83.0–89.6) | 98.9% (97.1–99.6) | 18.2% (10.7–29.1) | 16 (3.8%) | 79.0% |
| **ASCVD** | 81.8% (61.5–92.7) | 72.5% (66.8–77.5) | 98.0% (94.9–99.2) | 19.8% (12.9–29.1) | 22 (7.7%) | 61.2% |
| **No ASCVD** | 62.5% (30.6–86.3) | 92.9% (89.3–95.3) | 98.9% (96.9–99.6) | 19.2% (8.5–37.9) | 8 (2.6%) | 86.2% |
| **HF** | 94.4% (74.2–99.0) | 61.6% (53.6–69.1) | 98.9% (94.0–99.8) | 23.3% (15.1–34.2) | 18 (11.0%) | 52.9% |
| **No HF** | 50.0% (25.4–74.6) | 90.8% (87.6–93.2) | 98.4% (96.6–99.3) | 13.6% (6.4–26.7) | 12 (2.8%) | 79.3% |

<sup>a</sup>Data are presented as point estimate (95% CI), number (percentage), or percentage.
**Abbreviations:** ASCVD, atherosclerotic cardiovascular disease; HF, heart failure; LVH, left ventricular hypertrophy; LVSD, left ventricular systolic dysfunction; NNT, number needed to test; NPV, negative predictive value; PPV, positive predictive value; SHD, structural heart disease; T2D, type 2 diabetes.

Across the composite and individual phenotypes, the model maintained high specificity and NPV, while other metrics varied by phenotype (**Table 2**). For the broader SHD composite, sensitivity and specificity were 52.6% and 86.2%, respectively, with an NPV of 90.2% and a PPV of 42.9%. Detection was strongest for LVSD (sensitivity 86.7%, specificity 91.0%). For valvular disease, specificity stayed high (94.3%–94.5%), but sensitivity was lower (16.7% for severe and 21.5% for moderate or severe valvular disease). For severe LVH, sensitivity and specificity were 44.4% and 93.6%, respectively (**Table 2**).

### Number Needed to Test With and Without AI-ECG

AI-ECG–guided triage substantially lowered the NNT to identify one case relative to usual care (**Table 2**, **Figure 2B**). For severe SHD, the NNT fell from 19.6 to 5.1 (a 74.1% reduction) and for the broader SHD composite, from 6.1 to 2.3 (a 61.6% reduction). Across individual phenotypes, the relative NNT reduction was 87.4% for LVSD, 66.4% for severe valvular disease, 63.8% for moderate or severe valvular disease, and 84.3% for severe LVH (**Table 2**, **Figure 2B**). Of note, despite substantial reductions in NNT, NPV varied from 88.6% for moderate or severe valvular disease to 99.6% for LVSD (**Table 2**).

## DISCUSSION

In WATCH-SHD, we prospectively validated a noise-adapted deep learning model for detecting severe SHD from single-lead ECGs recorded on an Apple Watch through an end-to-end, HIPAA-compliant platform. Among 596 participants undergoing outpatient TTE, the model discriminated severe SHD well (AUROC, 0.841), with 76.7% sensitivity and 83.2% specificity, and performance held across the broader SHD composite and individual phenotypes. Used as a triage step before TTE, AI-ECG lowered the NNT to identify one case by more than 60% versus usual care across composite and individual outcomes—supporting wearable single-lead ECGs as a scalable screening modality for SHD.

To our knowledge, WATCH-SHD is the first prospective study to validate an AI-ECG approach for detecting a composite of clinically actionable SHD using real-world, point-of-care single-lead ECGs from a consumer wearable during routine care—extending prior work that focused largely on LVSD only.^21–23^ The advance matters because, even with strong discrimination, point-of-care screening is limited by PPV when the target phenotype is rare. Aggregating several actionable phenotypes into one composite label raises event prevalence, and therefore PPV, while keeping the result clinically meaningful for downstream testing and treatment.^17–19^

Because the prospective wearable AI-ECG literature has reported mainly on LVSD only, we used LVSD as the most direct benchmark. In WATCH-SHD, LVSD detection reached 86.7% sensitivity and 91.0% specificity, comparing favorably with a smartwatch-enabled Apple Watch study (75.0% sensitivity, 78.5% specificity; n=421),^21^ an ECG–enabled stethoscope study using the Eko DUO (82.7%, 79.9%; n=1050),^22^ and a six-lead portable ECG using the KardiaMobile 6L (83.4%, 88.7%; n=1635).^23^ These comparisons situate our LVSD performance within existing evidence, while our primary analyses extend evaluation to the composite SHD phenotype.

The sensitivity (76.7%) and specificity (83.2%) observed for detecting severe SHD support AI-ECG as a front-line triage test that flags patients for confirmatory imaging. Comparable operating characteristics are routinely accepted in cardiology for noninvasive tests—such as stress imaging for coronary artery disease—that guide selection for invasive coronary angiography.^31,32^ Here, the downstream test is TTE, itself noninvasive and low-risk, making AI-ECG well suited to raising the yield of echocardiography where imaging capacity is limited, and prioritization is needed.

False positives remain a central barrier to AI-enabled screening, since downstream testing and follow-up must stay acceptable in volume, cost, and patient burden.^33–36^ In WATCH-SHD, PPV was modest for severe SHD (19.7%) and higher for the broader composite (42.9%), tracking the composite’s greater prevalence. Because PPV depends on prevalence, the efficiency seen in this echocardiography-referred cohort may not carry over directly to lower-prevalence community settings. Beyond composite labeling, yield could be improved by applying AI-ECG in higher-risk populations—such as individuals with stage A heart failure,^37^ by combining its output with clinical risk features to prioritize imaging, or by using foundational AI-EHR models to surface high-risk individuals across a health system.^38^

Several limitations warrant mention. First, WATCH-SHD was conducted at a single echocardiography laboratory among patients already referred for outpatient TTE, which likely raised outcome prevalence and may overestimate PPV relative to community screening. Nonetheless, this design enabled rigorous prospective evaluation against a clinical reference standard and establishes real-world feasibility and operating characteristics that can now be tested in population-based settings. Second, SHD phenotypes were drawn from routine clinical TTE reports rather than core-laboratory adjudication and may be reader-dependent; even so, all measurements followed guideline-based practice and were interpreted by practicing cardiologists at a large academic center, supporting internal validity. Third, performance estimates for rarer phenotypes—severe valvular disease and severe LVH—rest on few events and are correspondingly imprecise, whereas the primary and composite endpoints most relevant to screening had more events and yielded stable, interpretable results. Fourth, performance reflects a single supervised Apple Watch workflow, and generalizability to unsupervised use or other wearables will require dedicated validation. Finally, we did not evaluate downstream clinical impact or health-system outcomes; these data provide the prospective foundation for pragmatic and interventional studies assessing implementation, risk-enriched deployment, and clinical benefit.

## CONCLUSION

In a prospective cohort undergoing outpatient echocardiography, a previously developed, noise-adapted AI-ECG identified SHD phenotypes from real-world single-lead ECGs recorded on an Apple Watch. Performance was consistent across the composite and individual phenotypes, substantially reducing the number of echocardiograms needed to identify one case, with deployment through an end-to-end, HIPAA-compliant platform supporting real-world feasibility. Together, these findings provide prospective evidence that wearable single-lead ECG–based screening could enable scalable identification of clinically actionable SHD, warranting evaluation in population-based and interventional studies.

## Acknowledgments

None.

## Data Availability Statement

The data from the Yale New Haven Health System represent protected health information. To protect patient privacy, the Yale Institutional Review Board does not allow the sharing of these data.

